# Perceived risk of type 2 diabetes: Using linked genomic, clinical and questionnaire data to understand the potential use of genetic risk tools in British South Asians

**DOI:** 10.1101/2024.03.01.24303599

**Authors:** Jing Hui Law, Daniel Stow, Sam Hodgson, Genes & Health Research Team, David A. van Heel, William G. Newman, Magda Osman, Sarah Finer

## Abstract

**Background:** Despite growing interest surrounding the integration of genetic risk tools such as polygenic risk scores (PRSs) into routine care for early disease identification and management, major questions remain about whether and how these tools are to be implemented at-scale. Many interventions have explored their use in encouraging the adoption of preventative health behaviours—yet existing evidence remains undetermined, limited by the focus on White European populations. The present study used structural equation modelling to explore genetic risk perceptions surrounding type 2 diabetes (T2D) in a sample of British Bangladeshi and British Pakistani volunteers—combining questionnaire data alongside genomic and clinical information to identify the characteristics of individuals who are likely to act on genetic risk information.

**Methods and findings:** We conducted this study with volunteers enrolled in Genes & Health—a large-scale (n > 60,000) study in the UK recruiting British Bangladeshi and British Pakistani volunteers from community and NHS settings. Eligible participants between the ages of 16 to 59 years were invited to complete a 15-minute questionnaire containing measures of genetic risk perceptions surrounding T2D, as well as intention to adopt health behaviours and that can prevent or delay T2D. Questionnaire responses were then integrated with participants’ genomic and clinical data available at Genes & Health to construct a model—characterising their mediating relationships in informing participants’ intention.

626 participants responded to the questionnaire (response rate = 17%, 37.70% aged 46 to 59 years, 62.62% female). Being between the ages of 46 to 59 years (β = 0.52, 95% CI [0.26, 0.79], p < 0.05), having greater self-reported perceived control over health (β = 0.41, 95% CI [0.26, 0.56], p < 0.05) and interest in genetic testing (β = 0.62, 95% CI [0.46, 0.78], p < 0.05) all had direct positive effects on participants’ intention. Household income showed an indirect effect on intention, mediated by interest in genetic testing, β = 0.24, 95% CI [0.12, 0.37]. Self-identified ethnicity also demonstrated indirect effects on intention via two mediating pathways—both involving participants’ actual T2D PRSs and self-reported family history of T2D (β = 0.03, 95% CI [0.02, 0.05] and β = 0.002, 95% CI [0.001, 0.01]).

**Conclusions:** Our results showed that older age, greater perceived control over health and interest in genetic testing are all predictive of participants’ likelihood of adopting preventative heath behaviours in response to genetic risk information about T2D. We also found evidence pointing to the roles that wider socio-demographic, clinical and familial variables can play in informing and mediating genetic risk perceptions. These findings should raise awareness about potential challenges to the equitable delivery and management of genetic risk tools—and strengthen calls for wider family- and system-level approaches that can help address potential health inequalities, as efforts surrounding the large-scale implementation of genomics into existing health systems continue to grow.

**Author summary:** *Why was this study done?:*

- Type 2 diabetes (T2D) disproportionately affects populations of South Asian ancestry.
- The integration of genetic risk tools such as polygenic risk scores (PRSs) into routine care has been widely discussed—and presents potential clinical utility in the early identification and management of T2D in British Bangladeshi and British Pakistani populations.
- Studies have explored the use of PRSs in shifting individuals’ genetic risk perceptions and/or encouraging the adoption of preventative health behaviours—yet existing evidence is limited by the focus on older and healthier White European populations.

*What did the researchers do and find?:*

- Combining questionnaire responses with genomic and clinical data in a sample of British Bangladeshi and British Pakistani volunteers, we applied structural equation modelling to analyse their mediating relationships—and to identify the characteristics of individuals who are likely to act on genetic risk information about T2D.
- Older participants in our sample reported greater levels of intention to adopt health behaviours that can prevent or delay T2D—however, most participants (34.5%) also indicated interest in finding out about genetic risk for T2D at younger ages.
- We found that relationships between participants’ actual and perceived risk for T2D were predominantly mediated by having first degree family member(s) with T2D history, compared to other clinical variables such as BMI or comorbidities.
- There were also mediating effects between participants’ self-reported household income and interest in genetic testing in predicting their likelihood of adopting preventative heath behaviours in response to genetic risk information about T2D.

*What do these findings mean?:*

- Upstream determinants and contextual factors such as socio-demographic characteristics and family history of disease can play important roles in leveraging the use of genetic testing for T2D in British Bangladeshi and British Pakistani populations.
- As efforts around the large-scale implementation of genomics into routine care continue to grow, future work should explore ways to integrate wider family- and system-level approaches that can help address potential health inequalities.
- It will also be important to consider how strategies can be tailored to younger age groups— given possible discordance between the age at which individuals would want to find out about genetic risk information, versus the age at which they would actually be willing to implement preventative lifestyle changes.

## Introduction

Type 2 diabetes (T2D) and its related complications have a disproportionately high prevalence and early onset among individuals of South Asian ancestry [1]. Emerging research has demonstrated that combining genetic risk tools such as polygenic risk scores (PRSs) with QDiabetes—a clinical risk model commonly used in the NHS—can improve the prediction of incident T2D in these populations [2]. Performance is especially enhanced for British Bangladeshi and British Pakistani populations at younger ages and lower body mass index (BMI)—who would otherwise have been considered healthy by QDiabetes alone [2]. Since individuals’ genetic liability to T2D is fixed and remains stable from conception, these findings demonstrate the unique benefit of tools such as PRSs to aid in early disease identification and management—prior to the development and accumulation of clinical and/or lifestyle-related risk factors that conventional clinical risk models rely on [3, 4].

At present, PRSs are extensively applied in discovery research—and there is growing interest around their clinical implementation on a population-wide basis [4-7]. Properly translated into clinical settings, improvements in predictive performance can present benefits such as further individualised screening for high-risk individuals and/or earlier referrals onto preventative care [8]. These efforts have been echoed by continuous calls for the wide-scale integration of genomics into routine care in England through the NHS [9-11]. Policies and strategies set out by the Genomic Medicine Service in 2022 describe priority areas such as delivering equitable genetic testing for cancer, rare, inherited and common diseases over the next 5 years—as well as the integration of genomics with other diagnostic and clinical data. As part of these initiatives, there is also a need to better understand the impact of providing individuals with personalised health and risk information using tools such as PRSs—generating evidence that can contribute to decisions on whether and how PRSs are to be implemented at-scale [11].

The clinical utility of PRSs in bringing about downstream population health benefits depend on two key factors: (1) that acting on disease risk can modify an individual’s health outcomes; and (2) that the individual informed of their risk may be willing and able to undertake the relevant preventative actions [12]. Many interventions have thus explored the use of genetic risk information in not only shifting individuals’ perceptions surrounding a disease—but also in encouraging the adoption of health behaviours that can prevent or delay disease onset [12-16]. However, current evidence surrounding cardiometabolic diseases is inconclusive—as their multifactorial aetiology and the need for sustained lifestyle changes can pose complex challenges [12, 16, 17]. Existing research is also limited by the focus on older and healthier White European populations—thus significant gaps remain surrounding how interventions should begin to address wider contextual factors and upstream determinants that may bring about different responses in diverse populations [17]. In particular, whether family experiences with common diseases can correspond to specific motivators for preventative health behaviours has been increasingly acknowledged—especially in light of emerging work on the complementary effects of family history information and genome-wide PRSs in capturing individuals’ risk across 24 common diseases [18]. There needs to be careful consideration around how these increasingly comprehensive ways to assess inherited disease risk can be translated in practice—and how broader influences of risk perceptions and/or health behaviours can be leveraged for the effective communication of genetic risk.

The broad aim of this study was to take a multidisciplinary methodological approach to study genetic risk perceptions surrounding T2D in a sample of British Bangladeshi and British Pakistani volunteers—exploring self-reported questionnaire data in the context of wider contextual factors, including participants’ actual genomic and clinical information. We undertook our study with volunteers enrolled in Genes & Health—a large-scale biobank in the UK which has recruited over 50,000 British Bangladeshi and British Pakistani participants from community and NHS settings [2, 19]. Combining the rich data resource in Genes & Health with a large-scale questionnaire on genetic risk perceptions with 626 volunteers, we used structural equation modelling (SEM) to characterise the mediating relationships between various socio-demographic, genomic, clinical and questionnaire variables—and to identify the characteristics of individuals who are likely to act on genetic risk information about T2D.

## Methods

### Design

Our integrated study design and analysis brought together multiple data sources in Genes & Health— investigating questionnaire-derived genetic risk perceptions alongside the genomic and clinical datasets available. Volunteers who were already enrolled in the biobank were invited to complete a cross-sectional, online-based questionnaire on genetic risk perceptions surrounding T2D via the online questionnaire platform REDCap. Questionnaire responses were then linked to genomic and clinical data via participants’ pseudonymised NHS numbers.

### Participants

#### Genes & Health

Genes & Health operates under ethical approval from the London South East National Research Ethics Committee (REC) and Health Research Authority (HRA), with Queen Mary University of London as Sponsor [19]. Details about the cohort have been described elsewhere [2, 7, 19]—and an overview of its recruitment process is included in S1 Appendix. In brief, British Bangladeshi and British Pakistani volunteers aged 16 years and above have donated saliva samples for DNA extraction and genetic tests, provided consent for researchers to access their electronic health records (EHRs), as well as consent to be recontacted (up to four times per year) for recall studies via separate ethics applications. The present study is linked to its original REC/HRA approvals (14/LO/1240). An application was first made to the Genes & Health Executive for internal review—and then submitted as an ethics amendment to REC/HRA. Approval was obtained on the 15^th^ of August 2022.

#### Inclusion criteria

To be eligible for the questionnaire, participants had to:

1. Be between the ages of 16 to 59 years;
2. Have no previous diagnosis of T1D or T2D in their primary care records; and
3. Have an email address and/or phone number registered with Genes & Health.

Volunteers meeting the above inclusion criteria were identified using their linked and pseudonymised demographic and health data stored in the Genes & Health Trusted Research Environment (TRE), as of the July 2022 data release. Details about our eligibility screening process are also presented in S1 Appendix.

### Measures

#### Questionnaire data

An overview of the questionnaire is presented in Table 1—with details of specific items and scoring methods described in S2 Appendix. Most of these questions were adapted from established and validated measures of genetic risk perceptions available in the literature (newly developed measures will be specifically indicated in S2 Appendix)—and then further refined and optimised via Patient and Public Involvement (PPI). Prior to recruitment, we conducted workshops and one-to-one PPI sessions with volunteers of Bangladeshi and Pakistani descent in the UK (including those not involved in Genes & Health) to test and develop the questionnaire iteratively. These sessions revolved predominantly around checking understandability and acceptability of the questionnaire within our target population—as well as ensuring that questions are culturally sensitive and relevant to participants’ understanding of T2D. Where required, bilingual staff at Genes & Health were involved in these PPI sessions to aid with translations between Bengali/Urdu and English. The questionnaire has also been extensively reviewed by members of the Genes & Health research team, as well as the Genes & Health Community Advisory Group—to help identify any potential difficulties or sensitivities with the questionnaire items, as well as to oversee the feasibility of the study.

**Table 1.** Overview of questionnaire items.

| Category | Items |
| --- | --- |
| Genetic risk perceptions | Knowledge of the genetic basis of T2D |
|  | Perceived risk for T2D |
|  | Interest in genetic testing |
|  | Perceived benefits of genetic testing |
|  | Perceived control |
| Familial variables | Known family members and/or close social contacts with T2D history |
|  | Family health behaviours |
| Outcome variables | (Primary) Intention to adopt health behaviours that can prevent or delay T2D, if a genetic test shows above-average risk for the condition |
|  | (Secondary) Interest in receiving an email about further online resources on health behaviours that can prevent or delay T2D |

#### Genomic data

Genotyping in Genes & Health was performed on Illumina Infinium Global Screening Array v3 with additional multi-disease variants. Variants with call rates < 0.99 and/or minor allele frequencies < 1% were excluded, as were single nucleotide polymorphisms with imputation quality scores < 0.3. We excluded individuals unlikely to have Pakistani or Bangladeshi ancestry on the basis of principal component 1 lying 3 or more standard deviations (SDs) from the self-reported mean. Imputation was performed using the TopMED-r2 panel. We estimated participants’ genetic risk for T2D using externally-derived scores published in the PGSCatalog as of April 2023 [20]. We selected the best-performing score, assessed by beta estimated from multivariate logistic regression models adjusted for age, sex, ancestry, and the first 20 genetic principal components; this outperformed scores developed within Genes & Health, without risk of overfitting. This was a European-ancestry score derived in 898,130 individuals [21]. The score was scaled to a normal distribution with a mean of 0 and SD of 1.

#### Clinical data

Clinical data extracted from participants’ EHRs included BMI and the presence of comorbidities. BMI was obtained from their primary or secondary care records up to December 2022—to capture values closest to the point of questionnaire recruitment. Comorbidities were extracted using SNOMED and ICD-10 codes—and with codelists generated as part of the NIHR AI MULTIPLY consortium [22]. A list of 22 physical and psychological health conditions were selected—guided by the NHS Quality and Outcomes Framework clinical and public health indicators for 2023/24 in England, based on evidence of health conditions that are likely to benefit from improved primary care [23]. This encompassed conditions such as asthma, atrial fibrillation and hypertension (full list in S3 Appendix).

#### Socio-demographic data

Questions about participants’ highest level of education, annual household income and the number of people living in their household were included towards the end of our questionnaire (S2 Appendix). These were adapted from UK Biobank material (available on the UK Biobank online resource centre). Additional demographic information—including self-identified sex and ethnicity— was extracted from participants’ Genes & Health Stage 1 recruitment questionnaires.

### Procedure

Sample size calculations for the questionnaire were based on the original paper from which the primary intention outcome was extracted [24]. Details are presented in S1 Appendix, alongside further information about our recruitment strategy. A stratified random sampling approach was taken to ensure balanced representation across age and sex (S1 Appendix). We first identified a total of 5,000 eligible volunteers. After filtering out those who have recently been recalled for other studies in Genes & Health, 4,955 questionnaire invitations were scheduled (Fig 1). These were sent out via email and/or text messages, with questionnaire links uniquely integrated—such that participants who received both email and text message invitations could only access and complete the questionnaire once. An overall bounce rate of approximately 26% was observed across the invitations. Of the 3,667 invitations successfully sent out, 725 individuals started the questionnaire and 99 respondents dropped out. A total of 626 complete responses were collected, giving a final response rate of 17% (626/3,667).

**Fig 1.**
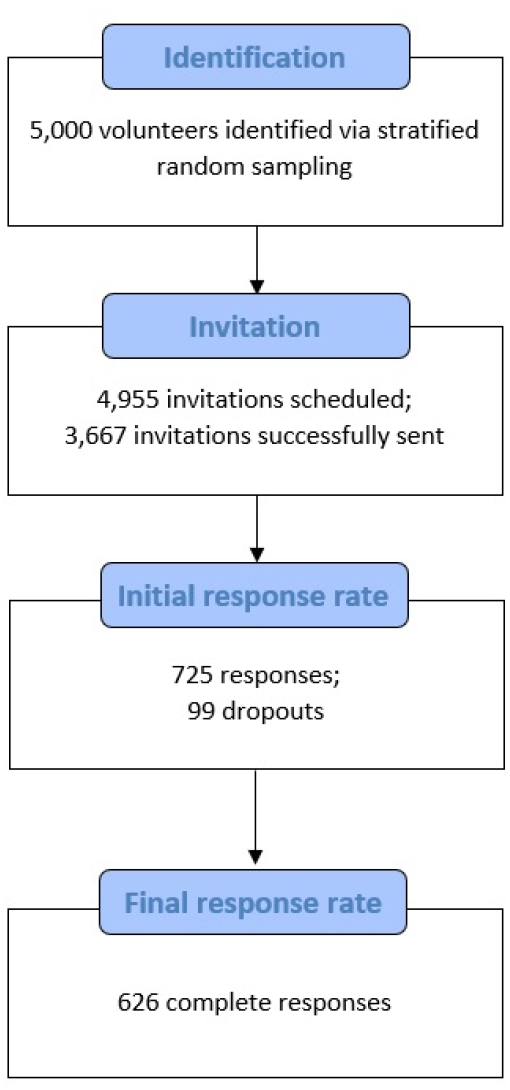
Flowchart illustrating participant recruitment.

On the REDCap landing page for the questionnaire, participants were first presented with a participant information sheet and consent form (S2 Appendix). Those who agreed to take part were then directed to answer further screening questions about age and diabetes history to fully ensure eligibility. Only participants who met these inclusion criteria could then proceed with the questionnaire. This took approximately 15 minutes to complete and participants were reimbursed with a £15 voucher. Recruitment closed in January 2023—and participants’ responses were securely exported from REDCap as an Excel data file, then uploaded back onto the TRE for data linkage and subsequent analysis.

### Data analysis

Descriptive analysis was first conducted to explore participant characteristics, as well as their genomic and clinical data obtained via linkage in Genes & Health. For the main analysis, SEM was performed using R packages “lavaan” [25] and “semTools” [26] to define and test a theoretical model incorporating all socio-demographic, genomic, clinical and questionnaire variables described above. SEM is a statistical procedure widely used to formalise and explore structural relationships between networks of variables and abstract constructs that cannot be directly measured or observed. Given the nature of constructs such as perceived control and intention involved in the current study, SEM allowed the opportunity to define and test mechanisms between these hypothetical constructs— estimating any direct or indirect relationships, whilst accounting for any potential measurement errors. As such, all multiple-item measures in the questionnaire were included as latent variables in the model—including participants’ knowledge of the genetic basis of T2D, interest in genetic testing, perceived control, perceived benefits of genetic testing, family health behaviours and the primary intention measure. Confirmatory factor analysis was conducted to define their measurement model and assess the validity of these latent variables (details in S4 Appendix). Cronbach’s alpha coefficients were also obtained to evaluate measurement reliability.

The main exogenous variable included in the final model was a binary variable for age—according to the age group with the largest *n* in our sample—to aid interpretation (46 to 59 years = 1; all other age groups = 0). Measurements of genetic risk perceptions, as well as the primary and secondary intention measures were included as endogenous variables. Other variables in the model included participants’ self-identified ethnicity, sex, as well as data extracted from their questionnaire responses on family history of T2D and family health behaviours. Socio-demographic variables such as annual household income and highest level of education were also coded for categories with the largest *n*—”Less than £18,000” and “College or University degree”, respectively. Number of comorbidities in the model was adapted to account for participants’ self-reported history of gestational diabetes and/or pre-diabetes from the questionnaire. There was missing data in participants’ T2D PRSs (n = 117; 18.69%) and BMI (n = 220; 35.14%) in our sample. Multiple imputation was thus applied alongside SEM, using R package “mice” [27] to create and analyse 30 imputed datasets—estimating missing values based on all other variables included in our model (e.g. age, sex and family history of T2D). All analysis was conducted using R version 4.2.1 [28].

## Results

Participant characteristics are presented below, alongside descriptive statistics for the genomic and clinical data obtained via linkage in Genes & Health (Table 2).

**Table 2.** Participant characteristics.

| **Participant characteristics (N = 626)** | **n (%) / Mean (SD)** |
| --- | --- |
| **Age group** |  |
| 16 to 25 years | 136 (21.73) |
| 26 to 35 years | 123 (19.65) |
| 36 to 45 years | 131 (20.93) |
| 46 to 59 years | 236 (37.70) |
| **Sex** |  |
| Male | 234 (37.38) |
| Female | 392 (62.62) |
| **Diabetes history** |  |
| Gestational diabetes | 39 (6.23) |
| Pre-diabetes | 63 (10.06) |
| No diabetes history | 524 (83.71) |
| **Number of people living in household** | 4.52 (1.69) |
| **Household income** |  |
| Less than £18,000 | 168 (26.84) |
| £18,000 to £30,999 | 114 (18.21) |
| £31,000 to £51,999 | 113 (18.05) |
| £52,000 to £100,000 | 78 (12.46) |
| Greater than £100,000 | 18 (2.88) |
| Do not know/prefer not to say | 135 (21.57) |
| **Education level** |  |
| College or University degree | 340 (54.31) |
| A levels/AS levels or equivalent | 113 (18.05) |
| O levels/GCSEs or equivalent | 57 (9.11) |
| CSEs or equivalent | 12 (1.92) |
| NVQ or HND or HNC or equivalent | 24 (3.83) |
| Other professional qualifications (e.g. nursing, teaching) | 13 (2.08) |
| None of the above/prefer not to say | 67 (10.70) |
| **T2D PRSs** | – 0.13 (1.00) |
| **BMI** | 25.92 (4.92) |
| **Number of comorbidities** | 1.42 (1.36) |

For the main analysis, diagonally weighted least squares estimation was used to fit the full model as shown in Fig 2. This hypothesised model demonstrated good fit across three of the imputed datasets, χ (323) = 622.90, *p* < 0.05, CFI = 0.97, TLI = 0.98 and RMSEA = 0.04, 90% CI [0.03, 0.04]. All pooled estimates are presented in Fig 2 below. Significant effects are denoted with solid lines (*p* < 0.05) and non-significant effects are denoted with dashed lines.

**Fig 2.**
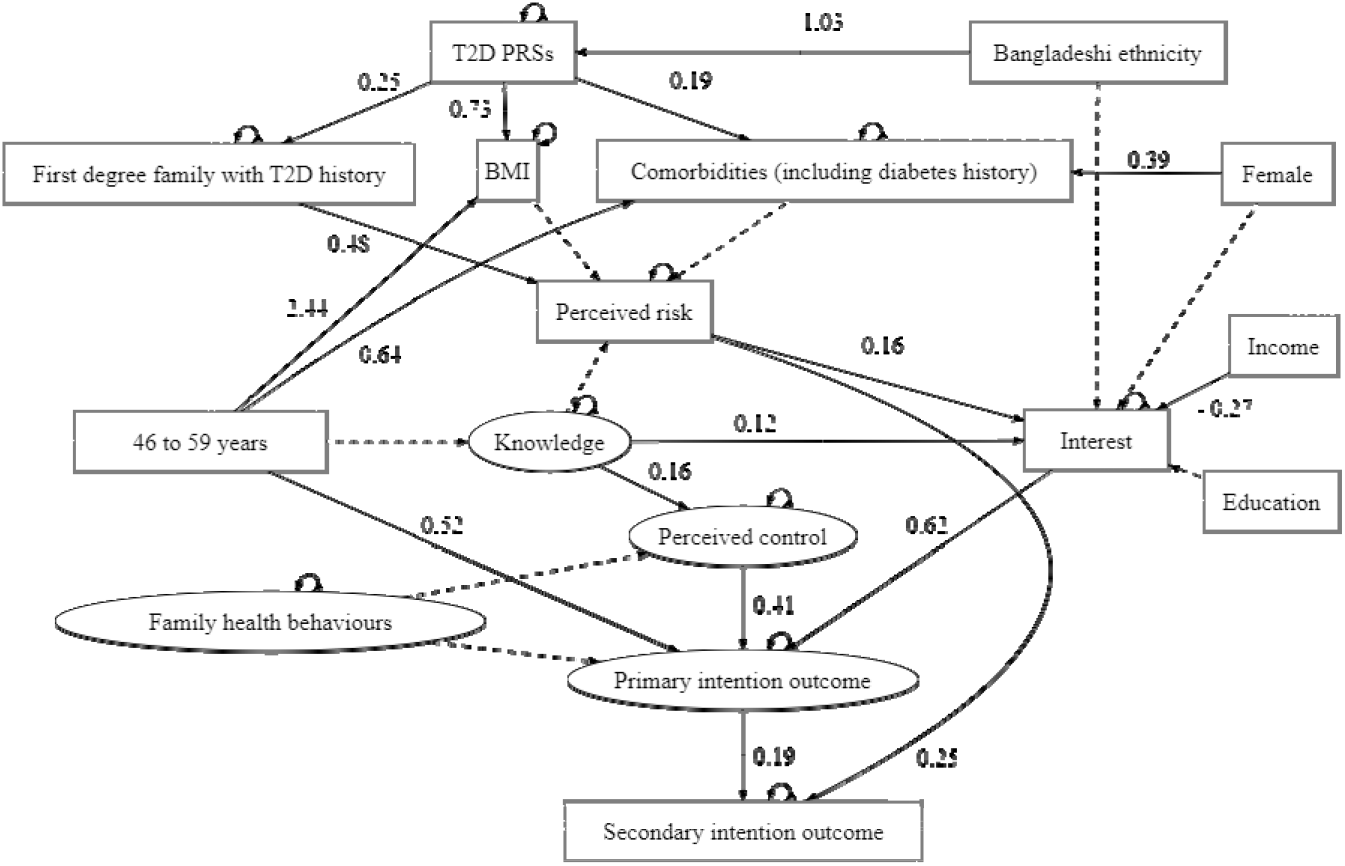
Hypothesised model for SEM.

### Direct effects

Being in the oldest age group (β = 0.52, 95% CI [0.26, 0.79], *p* < 0.05), having greater perceived control (β = 0.41, 95% CI [0.26, 0.56], *p* < 0.05) and greater interest in genetic testing (β = 0.62, 95% CI [0.46, 0.78], *p* < 0.05) all had positive direct effects on the primary outcome—i.e. participants’ self-reported intention to adopt health behaviours that can prevent or delay T2D, if a genetic test shows that they are at above-average risk for T2D. This, in turn, showed a positive direct effect (β = 0.19, 95% CI [0.10, 0.28], *p* < 0.05) on participants’ interest in receiving an email about further online resources on health behaviours that can prevent or delay T2D—a measure defined as the secondary outcome in this study.

In terms of other significant relationships between the genetic risk perceptions measured in the questionnaire, perceived risk had a positive direct effect on the secondary outcome (β = 0.25, 95% CI [0.15, 0.34], *p* < 0.05). Additionally, participants’ perceived control and interest in genetic testing were both predicted by their self-reported knowledge of the genetic basis of T2D—β = 0.16, 95% CI [0.06, 0.25], *p* < 0.05 and β = 0.12, 95% CI [0.02, 0.21], *p* < 0.05, respectively. Interest in genetic testing was significantly predicted by perceived risk (β = 0.16, 95% CI [0.06, 0.25], *p* < 0.05) and income level (β = – 0.27, 95% CI [– 0.53, – 0.01], *p* < 0.05). Perceived risk, in turn, was predicted by having first degree family member(s) with T2D history (β = 0.48, 95% CI [0.38, 0.57], *p* < 0.05). Hypothesised relationships between the other genomic and clinical variables included in our model indicated that participants’ T2D PRSs had direct effects on BMI (β = 0.73, 95% CI [0.26, 1.20], p < 0.05), comorbidities (β = 0.19, 95% CI [0.07, 0.31], p < 0.05) and having first degree family member(s) with T2D history (β = 0.25, 95% CI [0.15, 0.35], p < 0.05). T2D PRSs were, in turn, predicted by self-identified Bangladeshi ethnicity (β = 1.03, 95% CI [0.86, 1.19], p < 0.05). Comorbidities were predicted by sex (β = 0.33, 95% CI [0.07, 0.58], p < 0.05). Additionally, being in the oldest age group had direct effects on both BMI (β = 2.44, 95% CI [1.52, 3.36], p < 0.05) and comorbidities (β = 0.64, 95% CI [0.40, 0.89], p < 0.05).

### Mediation effects

In examining mediational pathways, Monte Carlo simulations were generated with 1,000 samples to yield robust 95% confidence intervals for the indirect effects, based on the estimated model parameters. Being in the oldest age group had a significant positive indirect effect on the secondary outcome, through its influence on the primary outcome, β = 0.10, 95% CI [0.04, 0.17]. Income level also had an indirect effect on the outcome measures, mediated by participants’ self-reported interest in genetic testing, β = 0.24, 95% CI [0.12, 0.37]. Additionally, participants’ self-identified Bangladeshi ethnicity had positive indirect effects on the questionnaire outcomes via two mediating pathways. Firstly, its effect on the secondary outcome was mediated by T2D PRSs, having first degree family member(s) with T2D history and perceived risk, β = 0.03, 95% CI [0.02, 0.05]. Secondly, this relationship was also mediated by T2D PRSs, having first degree family member(s) with T2D history, perceived risk, interest in genetic testing and the primary intention measure, β = 0.002, 95% CI [0.001, 0.01].

## Discussion

This study aimed to take a multidisciplinary methodological approach to investigate T2D genetic risk perceptions in British Bangladeshi and British Pakistani volunteers enrolled in Genes & Health— exploring questionnaire data in the context of participants’ actual genomic and clinical information. Combining the rich data resource in Genes & Health with a large-scale questionnaire with 626 volunteers, SEM was performed as the main analysis to define and test a theoretical model— incorporating various socio-demographic, genomic and clinical variables to characterise their mediating relationships alongside participants’ T2D genetic risk perceptions—and using this to identify the characteristics of individuals who are likely to act on genetic risk information about T2D.

Our results suggest that participants in the oldest age group (46 to 59 years) tended to report greater intention to adopt health behaviours that can prevent or delay T2D, if a genetic test were to show that they are at above-average risk for the condition. This ultimately fed into their interest in receiving an email about further online resources on T2D prevention—an outcome which was taken as a secondary measure of intention in the questionnaire. This finding is consistent with previous work suggesting that older populations usually demonstrate greater interest in seeking out disease risk information via genetic testing, compared to younger people [14, 15]. The saliency and relevance of health risks are often stronger in older adulthood—as this usually represents the stage of life where health concerns may first be emerging, yet there may still be time for behavioural changes to have a positive impact on health states [15]. Some studies have further reported that, compared to younger individuals, older adults usually demonstrate significantly stronger consistency between their self-reported intention—and the actual adoption of health behaviours at follow-up [29]. Such findings have been attributed to older adults having more established routines and habits—which may contribute to the regularity of their lifestyles and behaviours. Younger adults, on the other hand, are more likely to experience significant life adjustments (e.g. changes to living situations; forming new relationships)—which can explain the lack of alignment between their intention and behaviours [29]. It is worth noting, however, that when asked about their preferred age for genetic testing in our questionnaire, most participants (34.5%) still indicated that they would like to find out at younger ages—between 16 to 29—if they were genetically at risk of T2D. This suggests a certain level of discordance between the age at which individuals would want to find out about their risk, versus the age at which they would actually be ready—or willing—to implement preventative lifestyle changes.

Findings from our model also shed light on some of the roles that wider socio-demographic, clinical and familial variables can play in informing and mediating genetic risk perceptions surrounding T2D. For example, there were mediating effects between participants’ self-reported household income and interest in genetic testing in predicting outcomes on the questionnaire. Although overall effects were positive, that participants from lower income households in our sample reported less interest in genetic testing largely echo the gaps in health-related perceptions and behaviours that may exist between different socio-demographic backgrounds. This phenomenon should raise awareness about potential challenges to the equitable delivery and management of genetic risk tools. Whether due to a lack of awareness or understanding around the uses or purposes of genetic testing—or even perceived challenges or barriers to access health services—such differences point to the influence that characteristics linked to deprivation can have in driving health inequities. It also highlights the need for any efforts to integrate large-scale genetic testing on a population level to account for the social determinants of health—in order to fully maximise the utility and benefits of genetic risk tools. If PRSs are to be implemented at-scale, strategies will need to also target system-level factors— tackling the pathways and mechanisms driving potential health inequalities—so that individuals are not placed at further disadvantage. Furthermore, relationships between participants’ perceived and actual risk of T2D in our study sample were predominantly mediated by having first degree family member(s) with T2D history, even when compared to clinical factors such as BMI or comorbidities. This highlights the impact that previous experiences with T2D in the family context can have on how individuals think about their own risk—with further downstream effects on how they might readily react to genetic risk information. If relationships between participants ‘actual genetic risk status of T2D and their heightened sense of perceived risk is predominantly exerted through observing other family members with a history of T2D—even after accounting for other clinical variables—further work exploring how this can be leveraged in the communication of genetic risk is warranted, as there may be opportunities for interventions to tap into unique family experiences as drivers in encouraging preventative health behaviours. Other findings from our model showed that T2D PRSs are, in turn, predicted by being of self-identified Bangladeshi ethnicity. Additionally, female participants had more comorbidities recorded in their EHRs.

All these results lend support to the idea that the provision of genetic risk information should be combined with other forms of support to achieve goals of motivating preventative health behaviours more widely. There may be potential for educational interventions to be integrated alongside PRS delivery—to ensure that varying levels of understanding and/or interest in individuals of diverse socio-demographic backgrounds are addressed. Interventions can also be supplemented with further system-level services—incorporating elements such as environmental restructuring or social planning—to facilitate the translation from risk awareness into actual preventative action across diverse groups. This is especially important for individuals who may not necessarily have adequate resources and opportunities to implement lifestyle changes, despite being at high risk. Additionally, the potential role of family-based cascade testing may be an approach to consider for risk assessment. Unaffected family member(s) of patients who are already diagnosed with T2D likely already perceive themselves to be at high risk—thus baseline readiness to engage with preventative health behaviours may be higher than the general population—and offering genetic risk information about the condition may present unique benefits. However, how these relationships might play out in younger individuals at risk of T2D will need to be considered. Given PRSs often demonstrate the strongest clinical utility in younger populations, current findings around the discordance between the age at which individuals would want to find out about their genetic risk versus the age at which they would actually be willing to engage in preventative health behaviours are worth further exploration. It may be that, for younger populations to fully benefit, greater efforts combining system-level and environmental support services—integrating family, social and other resources—to fully engage preventative health behaviours will be needed to supplement the implementation of PRSs.

There were some limitations in this study—perhaps predominantly with regards to the representativeness of our questionnaire sample. It must be noted that respondents were recruited from a consented cohort, already involved in a large-scale genomics and health study. Although Genes & Health can broadly be considered representative of its background population [19], self-reported levels of genetic risk perceptions in the sample may not necessarily reflect the views of underlying British Bangladeshi and British Pakistani populations in the UK. Future work should aim to explore these issues more widely. Additionally, the original psychometric properties of some questionnaire measures that we have taken from the literature for this study have been affected following PPI procedures. However, these have been modified and refined according to item statistics (S4 Appendix)—with inadequate items that were limited in terms of validity or reliability removed before inclusion in the final model. On a similar note, the educational and income categories previously defined in UK Biobank might not necessarily reflect the same groups in Genes & Health. Descriptive statistics showed that whilst most participants in our sample reported an annual household income of less than £18,000, over half of the sample also reported being at least university- or college-educated. There may be other factors to consider here, such as participants’ immigration status, the countries where their educational qualifications were obtained—and whether such qualifications are equivalent to traditional definitions of being college- or university-educated in the UK. In future work, it may be that even seemingly objective socio-demographic measures also need to undergo a process of tailoring and refining to ensure that items are capturing the right categories in a diverse sample.

Nevertheless, this study has been able to provide some novel and unique insights into the perspectives surrounding genetic risk for T2D in an underrepresented population so disproportionately affected by the condition. It has also leveraged the rich genomic and clinical data available at Genes & Health to begin charting out the complexity of relationships underpinning these genetic risk perceptions. Taken together, our results point to the important roles that upstream determinants and contextual factors such as family history and household income can play in leveraging the use of genetic testing for T2D in British Bangladeshi and British Pakistani populations. As efforts surrounding the large-scale implementation of genomics into existing health systems continue to grow, future work should explore ways to integrate wider family- and system-level approaches that can help address potential health inequalities. It will also be important to consider how these strategies can be tailored to be applicable to younger age groups—given the discordance found in this study between the age at which individuals would want to find out about their genetic risk of T2D, versus the age at which they would actually be ready, or willing, to implement preventative lifestyle changes.

## Data Availability

All data produced in the present study are available upon reasonable request to the authors.

## Supporting information

S1 Appendix. Recruitment process in Genes & Health and the present study.

S2 Appendix. Questionnaire items.

S3 Appendix. Comorbidities included in analysis.

S4 Appendix. Results from confirmatory factor analysis.

## Acknowledgements

This work was made possible by funding from the Wellcome Trust for JHL through the doctoral training programme Health Data in Practice: Human-centred Science (Reference: 218584/Z/19/Z).

Genes & Health has recently been core-funded by Wellcome (WT102627, WT210561), the Medical Research Council (UK) (M009017, MR/X009777/1, MR/X009920/1), Higher Education Funding Council for England Catalyst, Barts Charity (845/1796), Health Data Research UK (for London substantive site), and research delivery support from the NHS National Institute for Health Research Clinical Research Network (North Thames). Genes & Health has recently been funded by Alnylam Pharmaceuticals, Genomics PLC; and a Life Sciences Industry Consortium of Astra Zeneca PLC, Bristol-Myers Squibb Company, GlaxoSmithKline Research and Development Limited, Maze Therapeutics Inc, Merck Sharp & Dohme LLC, Novo Nordisk A/S, Pfizer Inc, Takeda Development Centre Americas Inc.

We thank Social Action for Health, Centre of The Cell, members of our Community Advisory Group, and staff who have recruited and collected data from volunteers. We thank the NIHR National Biosample Centre (UK Biocentre), the Social Genetic & Developmental Psychiatry Centre (King’s College London), Wellcome Sanger Institute, and Broad Institute for sample processing, genotyping, sequencing and variant annotation.

We thank: Barts Health NHS Trust, NHS Clinical Commissioning Groups (City and Hackney, Waltham Forest, Tower Hamlets, Newham, Redbridge, Havering, Barking and Dagenham), East London NHS Foundation Trust, Bradford Teaching Hospitals NHS Foundation Trust, Public Health England (especially David Wyllie), Discovery Data Service/Endeavour Health Charitable Trust (especially David Stables), Voror Health Technologies Ltd (especially Sophie Don), NHS England (for what was NHS Digital)—for GDPR-compliant data sharing backed by individual written informed consent.

Most of all we thank all of the volunteers participating in Genes & Health.

Current Genes & Health Research Team (in alphabetical order by surname): Shaheen Akhtar, Mohammad Anwar, Elena Arciero, Omar Asgar, Samina Ashraf, Saeed Bidi, Gerome Breen, James Broster, Raymond Chung, David Collier, Charles J Curtis, Shabana Chaudhary, Megan Clinch, Grainne Colligan, Panos Deloukas, Ceri Durham, Faiza Durrani, Fabiola Eto, Sarah Finer, Joseph Gafton, Ana Angel, Chris Griffiths, Joanne Harvey, Teng Heng, Sam Hodgson, Qin Qin Huang, Matt Hurles, Karen A Hunt, Shapna Hussain, Kamrul Islam, Vivek Iyer, Benjamin M Jacobs, Ahsan Khan, Claudia Langenberg, Cath Lavery, Sang Hyuck Lee, Daniel MacArthur, Sidra Malik, Daniel Malawsky, Hilary Martin, Dan Mason, Rohini Mathur, Mohammed Bodrul Mazid, John McDermott, Caroline Morton, Bill Newman, Elizabeth Owor, Asma Qureshi, Shwetha Ramachandrappa, Mehru Raza, Jessry Russell, Nishat Safa, Miriam Samuel, Michael Simpson, John Solly, Marie Spreckley. Daniel Stow, Michael Taylor, Richard C Trembath, Karen Tricker, David A van Heel, Klaudia Walter, Caroline Winckley, Suzanne Wood, John Wright, Ishevanhu Zengeya, Julia Zöllner.

## Author Contributions

Conceptualisation: Jing Hui Law, Magda Osman, Sarah Finer.

Data curation: Jing Hui Law, Daniel Stow, Sam Hodgson, Genes & Health Research Team.

Formal analysis: Jing Hui Law.

Funding acquisition: Jing Hui Law.

Methodology: Jing Hui Law, Daniel Stow, Sam Hodgson, Genes & Health Research Team, Magda Osman, Sarah Finer.

Project administration: Jing Hui Law.

Supervision: David van Heel, Magda Osman, Sarah Finer.

Visualisation: Jing Hui Law.

Writing – original draft: Jing Hui Law.

Writing – review & editing: Jing Hui Law, Daniel Stow, Sam Hodgson, David van Heel, William G. Newman, Magda Osman, Sarah Finer.

## Notes

### Competing Interest Statement

The authors have declared no competing interest.

### Author Declarations

Genes & Health operates under ethical approval from the London South East National Research Ethics Committee and Health Research Authority (Reference: 14/LO/1240), with Queen Mary University of London as Sponsor.

