## Supporting information for "Perceived risk of type 2 diabetes: Using linked genomic, clinical and questionnaire data to understand the potential use of genetic risk tools in British South Asians"

### S1 Appendix. Recruitment process in Genes & Health and the present study.

Genes & Health takes a 2-stage recruitment process. In Stage 1, British Bangladeshi and British Pakistani individuals aged 16 and above—who are living in, working in or within reach of local communities in East London, Bradford and Manchester—are invited to participate. A convenience sampling approach is taken, with bilingual researchers recruiting volunteers from settings such as local mosques, libraries, GP surgeries and outpatient clinics. Stage 1 volunteers complete a brief baseline questionnaire, donate saliva samples for DNA extraction and genetic tests, provide consent for researchers to access their EHRs, as well as consent to be recontacted (up to four times per year) for Stage 2 recall studies.

Under these approvals, Stage 2 procedures in Genes & Health offer the opportunity to invite volunteers for more detailed study visits—e.g. for clinical assessment and/or the collection of biological samples, recall-by-genotype and/or phenotype—and also to develop trials-within-cohorts or sub-cohorts. These studies are, however, subject to separate ethics approvals—as well as volunteer acceptability and Genes & Health Community Advisory Group approvals. The present study takes place under such Stage 2 procedures.

Guided by the inclusion criteria set out for the questionnaire, we identified eligible volunteers using their linked and pseudonymised demographic and health data stored in Genes & Health, as of the July 2022 data release (S1 Fig).

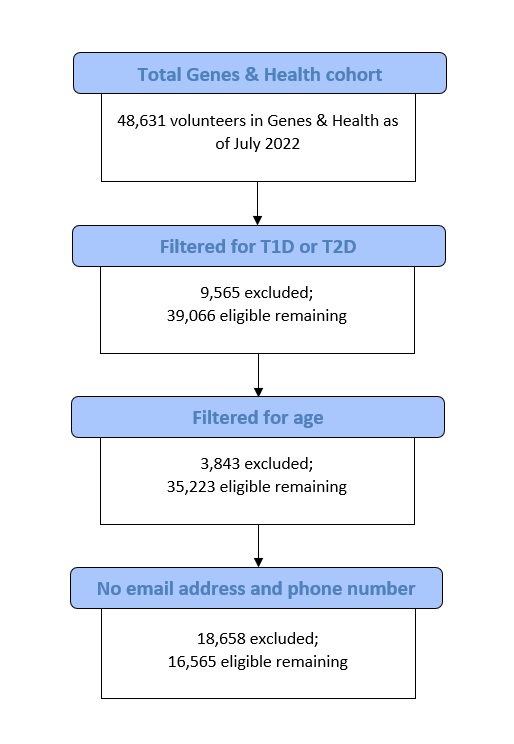

S1 Fig. Flowchart illustrating eligibility screening process.

Based on the original paper from which the primary intention outcome in our questionnaire was extracted, the authors estimated that a minimum sample size of 385 was needed to reach a power of 80%—with a confidence interval of 5%, a confidence level of 95% and SD of 0.05 [1]. To ensure balanced representation across four different age groups in the present study—i.e. 16 to 25 years; 26 to 35 years; 36 to 45 years and 46 to 59 years—it was agreed that at least 97 (385/4) participants in every age group was needed. Given the exploratory nature of the study, the final target of recruitment was set at approximately 120 participants per age group.

This was the first large-scale online questionnaire study conducted in Genes & Health—hence there were no available estimates surrounding possible response rates. In the first instance, we undertook a stratified random sampling approach to study recruitment. One thousand eligible volunteers were randomly selected according to each age group and sex strata—and then securely extracted from the Genes & Health TRE. In this initial phase, about 20% of all invitations could not be delivered to participants due to inactive email addresses and/or phone numbers. Of the invitations successfully sent, a total of 133 complete responses were collected—providing a response rate of approximately 17% (133/800). There were 51 (38%), 31 (23%), 33 (25%) and 18 (14%) responses from the youngest to oldest age groups, respectively.

Based on these figures, it was determined that an additional 4,000 eligible participants would be extracted from the TRE for the next phase of recruitment. Additionally, stratification was weighted for more participants in the older age groups—since the first phase of recruitment indicated progressively lower response rates from the youngest to oldest age groups. The breakdown of proportions in the participants extracted from the TRE for this second phase of recruitment was as follows—500 (12.5%), 750 (18.7%), 750 (18.7%) and 2000 (50%)—from the youngest to oldest age groups, respectively. Participants were again randomly selected within each age strata—and according to equal sex representation.

### S2 Appendix. Questionnaire items.

The following provides an overview of the items included in the study questionnaire. The complete questionnaire as presented on REDCap—including the participant information sheet and consent form—can be found further below.

#### Knowledge of the genetic basis of T2D

This was assessed using a newly established set of measures—rigorously developed in Genes & Health volunteers alongside a clinical geneticist, as part of a simultaneous study on genetic literacy conducted in Genes & Health. Feedback from the Genes & Health research team indicated this to be a measure relevant and important for the purposes of the present study. It contains five items, each measured on a five-point scale—and higher scores indicate higher levels of knowledge of the genetic basis of T2D.

#### Perceived risk for T2D

This was evaluated using a single-item measure, inviting participants to rate their thoughts on their personal risk of getting T2D in their lifetime on a four-point scale (almost no risk; a slight risk; a moderate risk; a high risk). It was adapted from a measure of perceived risk that has already been widely used in the literature examining genetic risk perceptions for common conditions, including T2D [2]. Higher scores on this measure reflect higher perceived risk.

#### Interest in genetic testing

This section began with a short excerpt explaining the nature and purpose of genetic tests in the context of T2D (details below). Participants were then asked to answer a range of questions about their interest in getting these tests themselves. Questions included the likelihood and importance for them to receive genetic tests for T2D—which were adapted from [3, 4]. Higher scores on these items indicate greater levels of interest in genetic testing for T2D. We included a follow up question asking participants who were interested in receiving genetic tests to report the age at which they would prefer to find out, if they had high genetic risk for T2D. Additionally, there was a separate item asking participants to report if they have ever personally paid for a private genetic test to find out about their heritage or ancestry. This was taken to further reflect participants’ general levels of interest in getting genetic tests.

#### Perceived benefits of genetic testing

This was measured using a set of six items adapted from [4]. The statements included both positive and negative outcomes of genetic testing—e.g. “Finding out about my genetic risk for developing type 2 diabetes would help me make important decisions about my health”; “Getting a genetic test would be a frightening or stressful experience for me”. Negative statements were reverse scored. All items were measured on a four-point scale (strongly disagree; disagree; agree; strongly agree)—and higher scores reflect greater perceived benefits of genetic testing.

#### Perceived control

Measurements of perceived control were adapted from a range of questions extracted from [2, 3]. There were four items, again covering both positive and negative statements—e.g. “Overall, I feel that I am able to control my health”; “If a genetic test tells me that I have an above-average risk for type 2 diabetes, then I would think that type 2 diabetes cannot be prevented”. Negative statements were reverse scored. Items were also measured on a four-point scale (strongly disagree; disagree; agree; strongly agree)—where higher scores reflect greater perceived control.

#### Familial variables

The next section of the questionnaire included a range of familial variables—including questions on participants’ known family members and/or close social contacts with T2D history. Here, traditional measures assessing family history information were expanded upon to include participants’ extended family members and/or close social contacts as potential options. These included aunts, uncles, cousins, as well as “unofficial” family members such as family friends—in acknowledgement of larger familial and social networks within South Asian communities that can play important roles in informing individuals’ conceptions of risk and disease [5-7]. This wider range of options was strongly supported by volunteers with whom our questionnaire was workshopped.

The next questions were in relation to household information and family health behaviours—the latter of which was adapted from a version of the UK Diabetes and Diet Questionnaire which has been optimised and validated in South Asians [8, 9]. Through our PPI sessions, a final measure containing eight items was finalised, encompassing a range of lifestyle factors such as diet and physical activity. Higher scores on these items reflect healthier family health behaviours (some statements were reverse scored).

#### Outcome variables

The primary outcome measure of intention was adapted from existing work on T2D risk perceptions in minoritised ethnic groups in the United States [1]. Three items measured on four-point scales were included to capture participants’ intention to adopt health behaviours that can prevent or delay T2D, if a genetic test shows that they are at above-average risk for T2D. These encompass participants’ likelihood of seeking advice from healthcare professionals, increasing physical activity, as well as improving dietary habits. Higher scores indicate stronger self-reported intention.

Additionally, a secondary measure of intention was included in the questionnaire—asking participants if they would like to receive an email about further online resources on health behaviours that can prevent or delay T2D. A positive response on this item was taken as a further indicator of participants’ willingness and readiness to engage in preventative health behaviours.

**DNA & Future Risk of Diabetes**

Thank you for your interest in this study!

Please read the information below carefully.

British Bangladeshis and British Pakistanis have some of the highest rates of type 2 diabetes in the UK—and you may be personally affected by the condition. Genes—which contain DNA that individuals inherit from their parents—can be linked to a person’s risk of health conditions such as type 2 diabetes. In the future, the NHS might be able to give people DNA/genetic information about their health, so they get an earlier warning of their type 2 diabetes risk. However, we don't know if people would want to have this information given to them—or if they know what this risk means for them.

This survey will be a first step to help us understand how British Bangladeshis and British Pakistanis think about their risk of type 2 diabetes in the DNA. For example, people may have different beliefs about the level of personal control they have over their health, after receiving DNA information. We want to study how these beliefs are affected by age and experience of type 2 diabetes (e.g. in family members). For example, if a person already has a close a family member affected by type 2 diabetes, they may react differently to DNA information about their own health. If you would like to take part in this survey, please tick the check box below. If you have any specific questions or concerns, please contact the Genes & Health research team at.

Thank you very much for your time.

☐ I confirm that I have read and understood the information above. I would like to take part in the study.

**Part 1 of 10**

**Welcome and thank you for agreeing to take part in this study about DNA & Future Risk of Diabetes!**

**Your responses will be stored securely and anonymously. Any information you give us will remain confidential and it will only be used for the purposes of this survey.**

**Please answer the questions below to check if you meet the criteria for this study:**

| Please select your age group (in years): | ☐ Below 16  ☐ 16 – 29  ☐ 30 – 39  ☐ 40 – 49  ☐ 50 – 59  ☐ 60 and above |
| --- | --- |
| Has a health professional ever told you that you have diabetes? | ☐ Yes – Type 1 diabetes  ☐ Yes – Type 2 diabetes  ☐ Yes – Gestational diabetes (diabetes during pregnancy)  ☐ Yes – Pre-diabetes  ☐ Yes – Diabetes but unsure what type  ☐ No  ☐ Prefer not to answer |

**Part 2 of 10**

**The questions below relate to your understanding about DNA and type 2 diabetes. Please rate how much you agree or disagree with each of the statements below:**

| I understand what DNA is. | ☐ Strongly disagree  ☐ Disagree  ☐ Agree  ☐ Strongly agree  ☐ Not sure |
| --- | --- |
| I understand how DNA is shared in families. | ☐ Strongly disagree  ☐ Disagree  ☐ Agree  ☐ Strongly agree  ☐ Not sure |
| I know that DNA can affect health. | ☐ Strongly disagree  ☐ Disagree  ☐ Agree  ☐ Strongly agree  ☐ Not sure |
| I know that DNA can affect disease. | ☐ Strongly disagree  ☐ Disagree  ☐ Agree  ☐ Strongly agree  ☐ Not sure |
| I think that changes in the DNA are linked to whether a person will get type 2 diabetes. | ☐ Strongly disagree  ☐ Disagree  ☐ Agree  ☐ Strongly agree  ☐ Not sure |
| Which of these sources of health advice do you trust? (You can select more than one answer) | ☐ GP, family doctor or nurse  ☐ Religious leader (e.g. Imam or Sheikh)  ☐ Alternative doctor (e.g. herbal practitioner,  homeopathic doctor or Ayurvedic doctor)  ☐ Friends  ☐ Family  ☐ Counsellor  ☐ Teacher  ☐ Youtube videos  ☐ Social media (e.g. Facebook, Instagram or Twitter posts)  ☐ None of the above |

**Part 3 of 10**

**The question below relates to your views about your personal risk for developing type 2 diabetes. Please answer the statement below:**

| What do you think your risk of getting type 2 diabetes in your lifetime is? | ☐ Almost no risk  ☐ A slight risk  ☐ A moderate risk  ☐ A high risk |
| --- | --- |

**Part 4 of 10**

**Please read the following explainer carefully:**

**Your DNA and genetics run in your family and bloodline. It can affect your risk of getting certain health conditions. There are now tests and tools that have been developed to estimate an individual's genetic risk for diseases such as type 2 diabetes:**

**These genetic tests can be done using a sample of your saliva (spit). They will be able to tell you whether you are at low or high genetic risk of developing type 2 diabetes in the future. Genetic information in your body cannot be changed, and therefore, your genetic risk will be fixed in your lifetime. There may be lifestyle changes you can make to potentially prevent or delay type 2 diabetes (e.g. diet and physical activity). However, your genetic risk may affect whether these lifestyle changes will work for you.**

**Based on this information, please answer the questions below, which are related to your interest in genetic tests:**

| How important is it for you to know if you have a genetic risk for type 2 diabetes? | ☐ Not at all important  ☐ Not important  ☐ Somewhat important  ☐ Extremely important |
| --- | --- |
| If you were offered a genetic test for type 2 diabetes for free, how likely is it that you would take the test? | ☐ Not likely at all  ☐ Not likely  ☐ Somewhat likely  ☐ Extremely likely |
| At what age would you like to learn if you have a genetic risk for type 2 diabetes? | ☐ I would not like to know if I have a genetic risk for type 2 diabetes  ☐ Below 16  ☐ 16 – 29  ☐ 30 – 39  ☐ 40 – 49  ☐ 50 – 59  ☐ 60 and above |
| Have you ever signed up for a personal genetic ancestry test (e.g. 23andme, Ancestry.com)? | ☐ Yes  ☐ No  ☐ Do not know  ☐ Prefer not to answer |

**Part 5 of 10**

**The questions below relate to your views about the outcomes and consequences of genetic testing for type 2 diabetes. Please rate how much you agree or disagree with each of the statements below:**

| Finding out about my genetic risk for developing type 2 diabetes would help me make important decisions about my health. | ☐ Strongly disagree  ☐ Disagree  ☐ Agree  ☐ Strongly agree |
| --- | --- |
| Getting a genetic test would be a frightening or stressful experience for me. | ☐ Strongly disagree  ☐ Disagree  ☐ Agree  ☐ Strongly agree |
| Getting a genetic test would be a frightening or stressful experience for my family and/or loved ones. | ☐ Strongly disagree  ☐ Disagree  ☐ Agree  ☐ Strongly agree |
| If a genetic test tells me that I have an above-average risk for type 2 diabetes, I think that health professionals can help support my health and well-being. | ☐ Strongly disagree  ☐ Disagree  ☐ Agree  ☐ Strongly agree |
| Finding out about my genetic risk for developing type 2 diabetes would be important for future generations in my family. | ☐ Strongly disagree  ☐ Disagree  ☐ Agree  ☐ Strongly agree |
| If a genetic test tells me that I have an above-average risk for type 2 diabetes, I am likely to experience fear, anxiety and/or depression. | ☐ Strongly disagree  ☐ Disagree  ☐ Agree  ☐ Strongly agree |

**Part 6 of 10**

**The questions below relate to your beliefs about the level of control you have over developing type 2 diabetes. Please rate how much you agree or disagree with each of the statements below:**

| Overall, I feel that I am able to control my health. | ☐ Strongly disagree  ☐ Disagree  ☐ Agree  ☐ Strongly agree |
| --- | --- |
| If I am going to get type 2 diabetes, I think that there is not much I can do about it. | ☐ Strongly disagree  ☐ Disagree  ☐ Agree  ☐ Strongly agree |
| I think that my personal health behaviours, such as diet and physical activity, can control my risks of getting type 2 diabetes. | ☐ Strongly disagree  ☐ Disagree  ☐ Agree  ☐ Strongly agree |
| If a genetic test tells me that I have an above-average risk for type 2 diabetes, then I would think that type 2 diabetes cannot be prevented. | ☐ Strongly disagree  ☐ Disagree  ☐ Agree  ☐ Strongly agree |

**Part 7 of 10**

**The question below relates to whether or not you have ever had a family member and/or close social contact diagnosed with type 2 diabetes. Please answer below:**

| In the people you have close social contact with in your life, do any of them have type 2 diabetes? (You can select more than one answer): | ☐ Mother  ☐ Father  ☐ Brother or sister  ☐ Any grandparents  ☐ Any aunts or uncles  ☐ Any cousins  ☐ Partner or spouse  ☐ Friend  ☐ Other – blood related  ☐ Other – not blood related  ☐ Prefer not to answer |
| --- | --- |

**Part 8 of 10**

**The questions below relate to the health behaviours of people living in your household. Please select your answers based on each of the statements below:**

| Including yourself, how many people are living together in your household (include those who usually live in the house such as students living away from home during term)? | ☐ Enter number: _____  ☐ Do not know  ☐ Prefer not to answer |
| --- | --- |
| People in my household work out, exercise, or participate in physical activity. | ☐ Never or very rarely  ☐ Once a week or less often  ☐ 2 – 4 times a week  ☐ 5 – 6 times a week  ☐ 1 – 2 times a day  ☐ 3 or more times a day  *Please note that physical activity is defined here as exercise carried out beyond daily work, for example running, going to the gym, walking, yoga.* |
| People in my household eat vegetables. Include fresh, tinned and frozen vegetables and pulses like lentils, chickpeas and kidney beans. | ☐ Never or very rarely  ☐ Once a week or less often  ☐ 2 – 4 times a week  ☐ 5 – 6 times a week  ☐ 1 – 2 times a day  ☐ 3 or more times a day |
| People in my household eat fruits. Include fresh, frozen, tinned and dried fruit. Do NOT count fruit juices. | ☐ Never  ☐ Less than half the time  ☐ About half the time  ☐ Most of the time  ☐ All of the time |
| People in my household cook with any of the following:   - butter - ghee - lard - coconut oil - palm oil | ☐ Never or very rarely  ☐ Once a week or less often  ☐ 2 – 4 times a week  ☐ 5 – 6 times a week  ☐ 1 – 2 times a day  ☐ 3 or more times a day |
| People in my household eat sugary foods such as:   - gulab jamun - mishti - halva - jalebi - rasmalai - sweets - biscuits - chocolate - cakes or cake rusks - sweet popcorn | ☐ Never or very rarely  ☐ Once a week or less often  ☐ 2 – 4 times a week  ☐ 5 – 6 times a week  ☐ 1 – 2 times a day  ☐ 3 or more times a day |
| People in my household drink sugary drinks such as:   - hot drinks with sugar (such as tea or coffee with sugar) - non-diet fizzy drinks - squashes - mixers - energy drinks - fruit juices - sweetened milk drinks - flavoured syrups | ☐ Never or very rarely  ☐ Once a week or less often  ☐ 2 – 4 times a week  ☐ 5 – 6 times a week  ☐ 1 – 2 times a day  ☐ 3 or more times a day |
| People in my household ask for snacks between meals such as:   - biscuits - chocolate - cakes - crisps - corn puffs - salted nuts - Bombay mix | ☐ Never or very rarely  ☐ Once a week or less often  ☐ 2 – 4 times a week  ☐ 5 – 6 times a week  ☐ 1 – 2 times a day  ☐ 3 or more times a day |
| When people in my household ask for unhealthy foods, other family members try to offer a healthy alternative. | ☐ Never  ☐ Less than half the time  ☐ About half the time  ☐ Most of the time  ☐ All of the time |

**Part 9 of 10**

**The questions below relate what you would do if a genetic test tells you that you have an above-average risk for type 2 diabetes. Please rate your answers based on each of the statements below:**

| If a genetic test tells me that I have an above-average risk for type 2 diabetes, I am likely to seek advice to prevent or delay type 2 diabetes. | ☐ Not likely at all  ☐ Not likely  ☐ Somewhat likely  ☐ Extremely likely |
| --- | --- |
| If a genetic test tells me that I have an above-average risk for type 2 diabetes, I am likely to seek advice from these sources (You can select more than one answer): | ☐ GP, family doctor or nurse  ☐ Religious leader (e.g. Imam or Sheikh)  ☐ Alternative doctor (e.g. herbal practitioner,  homeopathic doctor or Ayurvedic doctor)  ☐ Friends  ☐ Family  ☐ Counsellor  ☐ Teacher  ☐ Youtube videos  ☐ Social media (e.g. Facebook, Instagram or Twitter posts)  ☐ None of the above |
| If a genetic test tells me that I have an above-average risk for type 2 diabetes, I am likely to increase my physical activity to prevent or delay type 2 diabetes. | ☐ Not likely at all  ☐ Not likely  ☐ Somewhat likely  ☐ Extremely likely  *Please note that physical activity is defined here as exercise carried out beyond daily work, for example running, going to the gym, walking, yoga.* |
| If a genetic test tells me that I have an above-average risk for type 2 diabetes, I am likely to improve my dietary habits to prevent or delay type 2 diabetes. | ☐ Not likely at all  ☐ Not likely  ☐ Somewhat likely  ☐ Extremely likely |
| At the end of this survey, would you like to receive an email about some further online resources on health behaviours that can prevent or delay type 2 diabetes? | ☐ Yes  ☐ No |

**Part 10 of 10**

**There can be inequalities in type 2 diabetes risk and treatment according to the backgrounds**

**that people come from. We would like to ask you about your household income and**

**educational background so that we can understand these inequalities. For example, your**

**education may affect how well you understand information about DNA or type 2 diabetes, and**

**your income might affect whether you can follow a healthy diet or exercise programme.**

**These questions might be sensitive. Your responses will be stored securely and anonymously.**

**Any information you give us will remain confidential and it will only be used for the purposes**

**of this survey:**

| What is the average total income (before tax) received by your household? | ☐ Less than £18,000  ☐ £18,000 to £30,999  ☐ £31,000 to £51,999  ☐ £52,000 to £100,000  ☐ Greater than £100,000  ☐ Do not know  ☐ Prefer not to answer |
| --- | --- |
| Which of the following qualifications do you have? (You can select more than one answer): | ☐ College or University degree  ☐ A levels/AS levels or equivalent  ☐ O levels/GCSEs or equivalent  ☐ CSEs or equivalent  ☐ NVQ or HND or HNC or equivalent  ☐ Other professional qualifications: e.g. nursing, teaching  ☐ None of the above  ☐ Prefer not to answer |

### S3 Appendix. Comorbidities included in analysis.

Number of comorbidities in this study was defined as an index reflecting accumulation across the following conditions for each participant—guided by the NHS Quality and Outcomes Framework clinical and public health indicators for 2023/24 in England (listed in alphabetical order):

1. Anxiety
2. Asthma
3. Atrial fibrillation
4. Bipolar affective disorder and mania
5. Cancer
6. Chronic kidney disease
7. Chronic obstructive pulmonary disease
8. Coronary heart disease
9. Dementia
10. Depression
11. Epilepsy
12. Heart failure
13. Hypertension
14. Learning disabilities
15. Obesity
16. Osteoporosis
17. Peripheral arterial disease
18. Polycystic ovary syndrome *
19. Rheumatoid arthritis
20. Schizophrenia and other psychoses
21. Smoking status
22. Stroke

* Polycystic ovary syndrome is not an NHS Quality and Outcomes Framework clinical or public health indicator. However, it was included in this list, given its role as a known significant risk factor for T2D.

### S4 Appendix. Results from confirmatory factor analysis.

Confirmatory factor analysis was conducted to assess the validity of the latent variables included in our model. Cronbach’s alpha coefficients were also obtained to evaluate measurement reliability for these constructs. An initial measurement model demonstrated acceptable fit, χ^2^(362) = 1680.75, *p* < 0.05, CFI = 0.77, TLI = 0.74 and RMSEA = 0.08, 90% CI [0.07, 0.08] (S2 Fig).

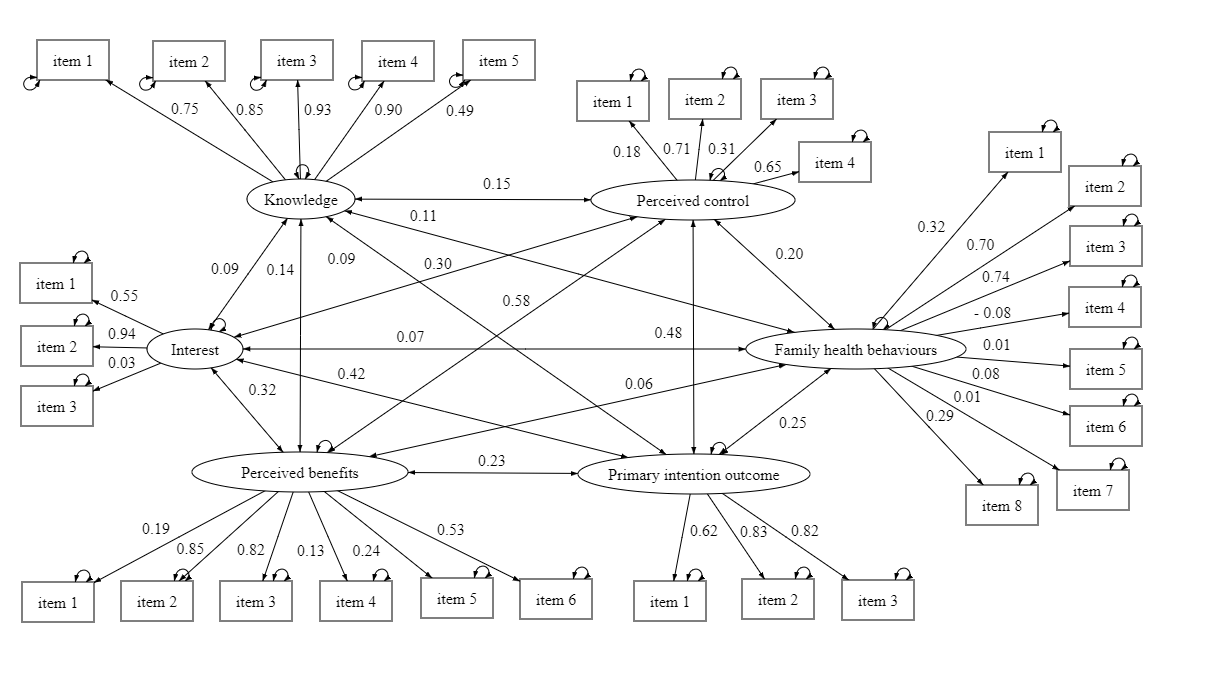

S2 Fig. Initial measurement model for latent variables.

For knowledge of the genetic basis of T2D, factor loadings were significant for all five items on the construct. Cronbach’s alpha coefficient was 0.89, suggesting good internal consistency. The measure was thus retained for analysis in the full model.

Interest in genetic testing was reduced into a one-item construct based on the item in the questionnaire with the highest factor loading and also deemed most theoretically relevant to the current study—“If you were offered a genetic test for type 2 diabetes for free, how likely is it that you would take the test?”. This was included in the final model as an observed variable.

For both perceived control and perceived benefits of genetic testing, factor loadings varied considerably within each construct—and internal consistency ranged from a poor 0.52 to 0.66. Due to some overlap between the content of questions in these two constructs, exploratory factor analysis was applied to a subset of the questionnaire data to review the dimensionality of these two sets of questions. Fitting a one-factor model showed that five items across these measures had factor loadings of over 0.30 (S1 Table). These items were mostly related to fear and anxiety over potentially negative outcomes of genetic testing—particularly a sense of loss of control over T2D prevention (S1 Table). A newly defined measurement model—combining this subset of five items—was then tested for a single measure of perceived control to be used in place of the two original measures. Cronbach’s alpha coefficient was 0.77 for this new measure, indicating improved internal consistency.

|  | Factor loading | Communality |
| --- | --- | --- |
| Getting a genetic test would be a frightening or stressful experience for me. | 0.81 | 0.65 |
| Getting a genetic test would be a frightening or stressful experience for my family and/or loved ones. | 0.76 | 0.58 |
| If a genetic test tells me that I have an above-average risk for type 2 diabetes, I am likely to experience fear, anxiety and/or depression. | 0.58 | 0.34 |
| If I am going to get type 2 diabetes, I think that there is not much I can do about it. | 0.48 | 0.23 |
| If a genetic test tells me that I have an above-average risk for type 2 diabetes, then I would think that type 2 diabetes cannot be prevented. | 0.55 | 0.30 |

S1 Table. Results from the exploratory factor analysis for perceived control and perceived benefits of genetic testing.

For the originally defined family health behaviours construct, factor loadings also varied considerably—and internal consistency was a poor 0.54. Exploratory factor analysis was similarly applied to a subset of the questionnaire data to review the dimensionality of this construct. Fitting a one-factor structure showed that four items relating to dietary habits in the family environment had factor loadings of over 0.30. Cronbach’s alpha coefficient for this four-item measure was, however, still unsatisfactory—at 0.58—and item statistics indicated that dropping the final item on the measure would improve reliability. Only three items were therefore included in the newly defined measurement of family health behaviours for the final model (S2 Table). Cronbach’s alpha coefficient was 0.70, suggesting good internal consistency.

|  | Factor loading | Communality |
| --- | --- | --- |
| People in my household eat sugary foods such as:   - gulab jamun - mishti - halva - jalebi - rasmalai - sweets - biscuits - chocolate - cakes or cake rusks - sweet popcorn | 0.60 | 0.36 |
| People in my household drink sugary drinks such as:   - hot drinks with sugar (such as tea or coffee with sugar) - non-diet fizzy drinks - squashes - mixers - energy drinks - fruit juices - sweetened milk drinks - flavoured syrups | 0.63 | 0.40 |
| People in my household ask for snacks between meals such as:   - biscuits - chocolate cakes - crisps - corn puffs - salted nuts - Bombay mix | 0.72 | 0.51 |

S2 Table. Results from the exploratory factor analysis for family health behaviours.

For the primary intention measure, factor loadings were significant for all three items on the construct. Cronbach’s alpha coefficient was 0.79, suggesting good internal consistency. The three-item measure was therefore retained for interpretation in the final model.

A revised model incorporating all the above changes was tested—and this demonstrated a slightly better fit than the initial model, χ^2^(98) = 254.54, *p* < 0.05, CFI = 0.92, TLI = 0.91 and RMSEA = 0.07, 90% CI [0.06, 0.08]. Modification indices were subsequently examined to identify further improvements—and these suggested the inclusion of correlated uniqueness between some of the items on the knowledge of the genetic basis of T2D scale, as well as on the newly defined measure of perceived control. The final measurement model showed a good fit, χ^2^(95) = 166.46, *p* < 0.05, CFI = 0.98, TLI = 0.98 and RMSEA = 0.04, 90% CI [0.03, 0.04]. This is presented in S3 Fig below, alongside all standardised factor loadings.

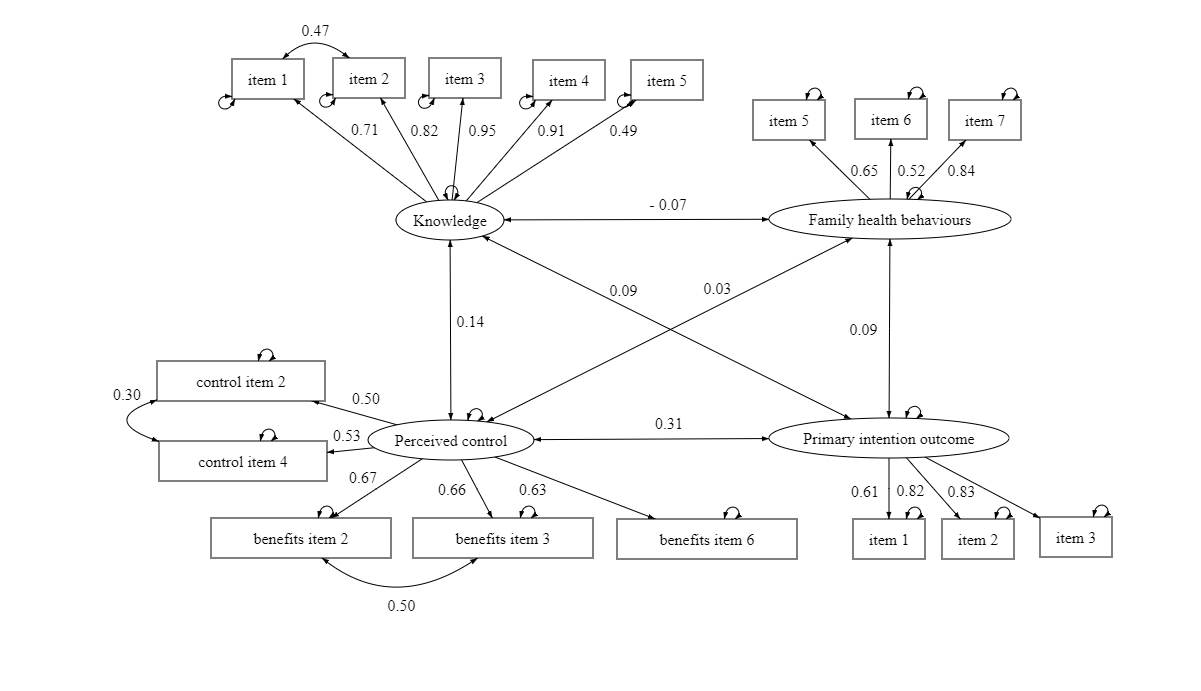

S3 Fig. Final measurement model for latent variables.
